# Indexing Cerebrovascular Health Using Transcranial Doppler Ultrasound

**DOI:** 10.1101/2020.04.20.20073197

**Authors:** R. Ghorbani Afkhami, R. Wong, S. Ramadan, F. R. Walker, S. J. Johnson

## Abstract

Transcranial Doppler (TCD) blood flow velocity has been extensively used in biomedical research as it provides a cost-effective and relatively simple approach to assess changes in cerebral blood flow dynamics and track cerebrovascular health status. In this paper, we introduce a new TCD based timing index, TI_tcd_, as an indicator of vascular stiffening and vascular health. We investigate the correlations of the new index and the existing indices, namely the pulsatility index (PI_t_cd) and the augmentation index (AI_TCD_), with age, cardiorespiratory fitness (CRF) and magnetic resonance imaging (MRI) blood flow pulsatility index (PI_mri_). Notably, the new TI_tcd_ index showed stronger correlations with CRF (*r* = −0.79) and PImri (*r* = 0.53) compared to AI_tcd_ (*r* = −0.65 with CRF and no significant correlation with PImri) and PI_TCD_ (no significant correlations with CRF or PImri) and similar correlations with age as AI_tcd_. The clearer relationship of the proposed timing index with vascular aging factors represented by cerebrovascular resistance (CVR) suggests its utility as an early indicator of vascular stiffening.

## Introduction

Changes in vascular stiffness are important indicators for vascular health independent of blood pressure [1] and the American Heart Association recommends the clinical measurement of vascular stiffness to predict future cardiovascular disease risk [1]. Vessel compliance describes the process of localized distension of the vascular wall in response to the pulsatile delivery of blood driven by the contraction of the heart. A stiff vessel is one which has lost its compliance. In a healthy vascular system, the compliant vessels act to smooth out the pressure waveform coming from the heart and reduce the peak pressure experienced by more distal vessels. Conversely, a loss of compliance (i.e. increased stiffening), results in increased pressure due to less efficient smoothing of the pressure waveform. This increase in pressure can compound other causes of elevated pressure such as vascular resistance caused by vessel narrowing.

As vessels lose their compliance, the speed of the pressure wave, called its pulse wave velocity (PWV), increases. PWV is measured by the time taken for the pressure wave, caused by the ejection of blood from the heart, to move down a given length of vessel. At a local level, when the pressure wave reaches a vessel bifurcation, a part of the wave is reflected towards the heart. Due to its fast speed the reflected wave can arrive back sufficiently close in time to the original wave front to augment the peak pressure exerted on vessels proximal to the recombination point. As a result of this process, small arteries, arterioles and capillaries face higher pressure and flow pulsations in each cardiac cycle. Over time these higher pressures can result in tearing of their endothelial and smooth muscle cells, producing a condition known as small vessel disease (SVD) [2,3]. SVD is especially important in the brain leading to cognitive decline, dementia, age-related disabilities and stroke [4].

Due to the well documented association between changes in compliance and serious brain pathology, there has been considerable research into techniques that measure changes in cerebrovascular compliance. Currently, the most common methodology to assess changes in the cerebrovascular compliance is Transcranial Doppler (TCD) ultrasound. TCD is a non-invasive technique that has been preferred because it is considered to be safe, cost-effective and relatively fast to perform. TCD is based on the “Doppler effect” according to which the ultrasound waves reflected back from the moving blood cells within the vessels have a different frequency from the emitted waves and this difference in frequency is directly proportional to the speed of those blood cells [5]. TCD is recognized to have excellent temporal resolution, making it ideal to capture information on blood flow dynamics.

Importantly, TCD can not measure compliance directly. However, several indirect indices have been developed including the pulsatility index (PI) and the augmentation index (AI) which are considered to provide reasonable approximations. The pulsatility index is the most commonly reported indirect measure of vascular health in the literature. PI_TCD_, first defined by [6], computes the peak to peak height of the flow velocity waveform divided by the mean flow velocity. Consistent with the idea that PI_TCD_ indexes vascular health, it has been shown to correlate robustly with aging [7-9] and aortic PWV [8]. Studies have reported correlations between PI_TCD_ and white matter disease [10,11], diabetes mellitus [12] and dementia [13].

There have been different interpretations of which particular vessel properties influence PI_TCD_ measurements. Some have proposed that PI_TCD_ measures cerebrovascular resistance (CRV) at the reflection site [11,13–16], while other studies have reported inconsistent results [17–20]. Following the discovery of vascular compliance, it has been hypothesized that as the vessels lose their compliance and become stiffer peak systolic (maximum) flow velocity increases and end-diastolic (minimum) flow velocity decreases which elevates calculated PI_TCD_ values [10]. More recently PI_TCD_ has been modeled as combination of resistance, compliance, PWV, and heart rate [7]. Consequently, PI_TCD_ is now often reported as an index of cerebrovascular health in general.

The second commonly used index of compliance is the augmentation index. AI was initially defined for the pressure waveform [21] and has been correlated with vascular aging and aortic PWV [22]. AI captures the reflected wave augmentation of the peak pressure by calculating the ratio of the reflected wave height to the systolic wave height with higher values said to result from earlier arrival of the reflected wave due to stiffer arteries. The AI index has been applied to TCD flow velocity waveforms in several studies [23,24] and positive and significant correlation between middle cerebral arterial (MCA) AI_TCD_ and age has been reported, *r* = 0.54 *(n =* 286) [25]. However, several studies have questioned the reliability of AI as an indicator of vascular compliance [26–28]. Studies have shown that AI also correlates with numerous biological factors not directly related to vessel properties including sex, heart rate, food intake, hydration status, height, weight and body composition [27].

An alternative TCD index to PI and AI that may provide a more direct measure of vessel health is to use a time-based index which directly measures the timing of the reflected wave. Evidence in an analysis of pressure waves suggests that a timing index can more accurately reflect vascular properties as it has a clearer relationship with vascular resistance, vascular compliance and PWV [29] and may be less susceptible to being influenced by biological factors that are independent of the vessel properties. In this paper we define a TCD timing index, TI_tcd_, as the inverse of the time between the systolic peak and the reflected waveform peak in the TCD signal. Thus, ti_tcd_ will track the timing of the blood waves in a similar way to PWV, the gold standard for vascular stiffness in central arteries. After introducing the new timing index, we investigate the correlation of the three TCD indices, PI, AI and TI, with other accepted measures of vascular aging such as magnetic resonance imaging (MRI) blood flow pulsatility index, age and cardiorespiratory fitness (CRF).

## Materials and Methods

Data from two different sets of experiments have been used in this study. Experiment one is a new study including Doppler, MRI and CRF whereas the second experiment uses Doppler recordings from an existing dataset.

### Experiment 1

Data from 38 adult volunteers (23 female and 14 male, age range = 24-66 years, mean age = 41.7 years) were recorded, which we will refer to as dataset D1. Participants were recruited from the local Novocastrian community, and signed an informed consent prior to any assessment. The study protocol is approved by the University of Newcastle Human Research Ethics Committee and registered in the Australian New Zealand Clinical Trials Registry (ACTRN12619000144112). Height, weight, age, gender and resting heart rate were recorded and the participants were asked to complete a physical activity questionnaire. Data acquisition was carried out over two imaging sessions on two consecutive days for each participant. Participants were asked to refrain from caffeine consumption before their scans. However, it was not an exclusion criteria in the study.

TCD ultrasound (DopplerBox X; Compumedics DWL, Singen, Germany) at the sampling rate of 100 samples per second was used to record cerebral blood flow velocity from the right and left middle cerebral arteries. The participants wore a headpiece with bilateral Doppler probes which stayed in place for the 300 seconds of resting-state recording. In this session, the resting-state heart rate was measured using OMRON HEM-7320 heart rate monitor device. Participants were in sitting position approximately five minutes during the headpiece setup, before the recording took place. During TCD recording, heart rate was measured once per minute three times. The three heart rate measurements were averaged later and used as the resting-state heart rate.

The participants were also scanned on a 3T MRI scanner (Magnetom Prisma, Siemens Healthineers, Erlangen, Germany), equipped with 64-channel receive only head coil, while a standard built in dual channel body coil was used for RF transmission. Blood flow was quantified using phase contrast flow quantification sequence (TR = 26.5ms, TE = 6.9ms, slice thickness = 5mm, matrix 256 × 256). A single excitation with the velocity encoding value 120 cm/s was used to quantify blood flow in middle cerebral arteries [30]. The TCD and MRI scan sessions were held on two consecutive days.

Two subjects did not have usable Doppler recordings and were excluded from further analysis. A further three did not have MRI data and were excluded from the Doppler versus MRI comparison.

### Experiment 2

This experiment was carried out using previously recorded TCD experiments examining cognitive performance with partial data being published previously in [31,32]. The aim of using this dataset, which we will refer to as D2, is to uncover the relationship of TCD indices with age in a population with wider age range including older participants as consistently lower correlations have been reported for PI_TCD_ in narrower and younger population age ranges [8,9, 12,33–35]. D2 has 55 subjects (34 female and 21 male, age range = 21-80 years, mean age = 45.8 years). The data was recorded with the same device as experiment one. Recording time was shorter than experiment one and different for subjects. Data was inspected manually for each subject and a 10-second-long segment with the highest quality was selected for further analysis. During manual inspection 3 subjects were excluded from the initial data as peak detection was not possible due to noise levels.

### Calculated Indices

*PI_TCD_:* Indices were calculated for both experiments. PI_TCD_ is a device output with values averaged per subject. The device calculates PI_TCD_ as defined by [6] i.e.,

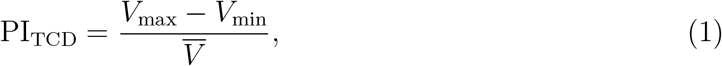

where *V*_max_, *V*_min_ and 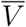 are maximum, minimum and mean flow velocities, respectively. An alternative definition, 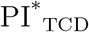, uses *V*_sys_ − *V*_dia_ in the numerator where *V*_sys_ and *V*_dia_ rare peak systolic and end-diastolic velocities, respectively. The two definitions report the same value in most cases. The difference arises when *V*_sys_ and *V*_max_ have different values. See Fig. 1. B for an example. This is the case in older populations and in stiffer arteries where the forward and the reflected waves meet early and *V*_sys_ hides on an inflection point whereas *V*_max_ becomes the reflected wave peak, *V*_refl_, see Fig. 1.B. Nonetheless, we compared PI_TCD_ values given by the device with manually calculated 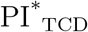 values for our experimental data and there was no notable difference. Therefore, only PI_TCD_ values directly calculated by the device are reported in this paper.

**Figure 1:**
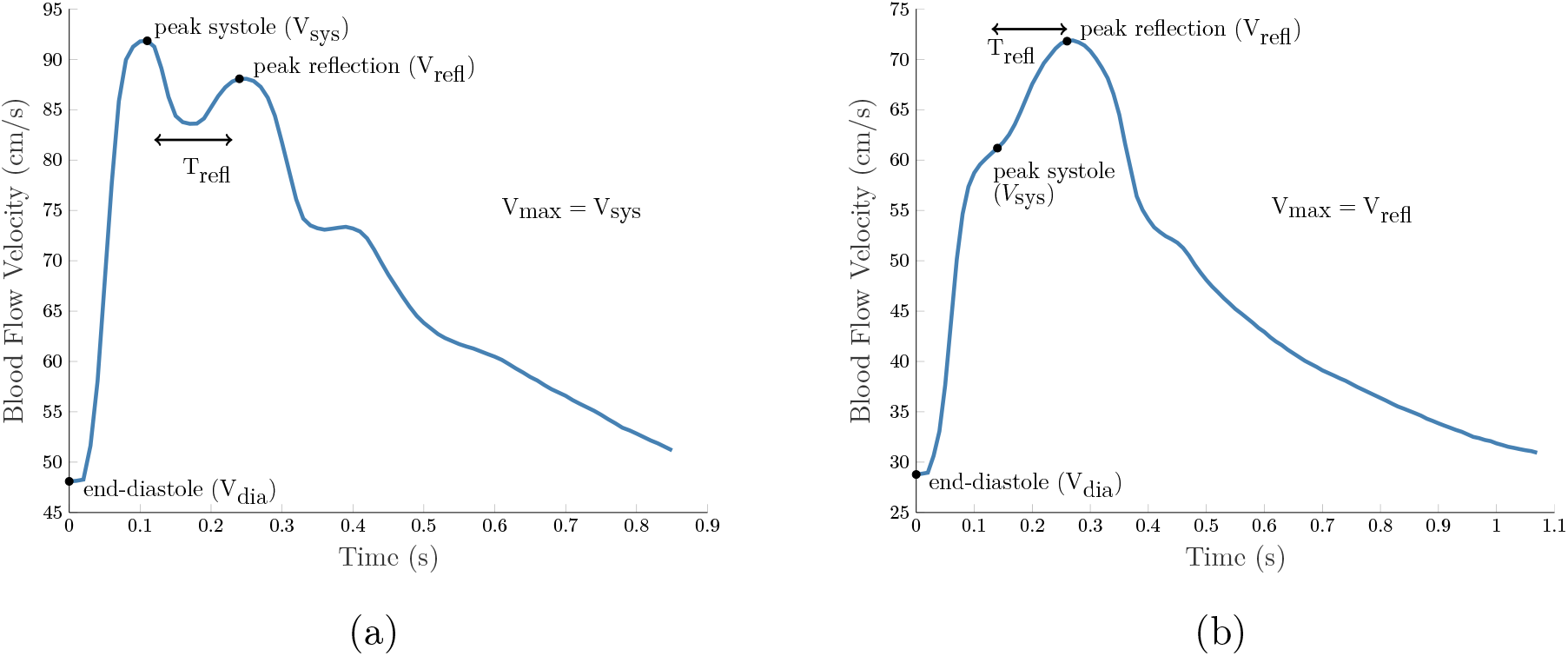
Two different TCD flow velocity samples from D1. (A) Averaged MCA, subject #22, age 25 years (B) Averaged MCA, subject #17, age 66 years

*AI_TCD_:* In order to calculate the augmentation index, first end-diastolic points were extracted automatically using Matlab by finding all the end-diastolic points (minimum flow velocity points) in each 300-second-long recorded signal for experiment one and 10-second-long signal for experiment two. The end-diastolic points were used to determine each cardiac cycle which were then averaged producing a short single-heartbeat long waveform per vessel of measurement. Next, the systolic peak, *V*_sys_, and the reflected peak, *V*_refl_ were located manually on the waveform and AI_TCD_ was calculated as

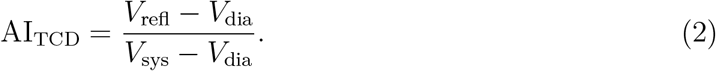

ai_tcd_ values were averaged across both MCAs, providing a single number per subject.

*pi_mri_:* The MRI pulsatility index, is defined similar to the TCD pulsatility index as [36,37]:

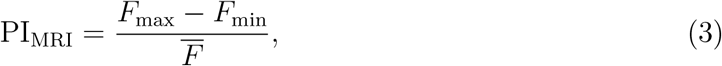

where *F* is the blood flow (in ml/sec) and the bar sign indicates the mean value. MRI scanner software (Siemens Syngo) was used to quantify flow values by placing a region of interest around the middle cerebral artery. The flow values for every repetition time, TR, produced a waveform for the length of the heart beat for which the maximum, minimum and the mean values were used to calculate PI_MRI_ following the method in [36]. Unlike TCD, for which only flow velocity is available, MRI measures of PI consider the flow (which combines flow velocity with vascular cross-sectional area) and is more closely tied to the definition of compliance. From *n* = 33 subjects of D1, PI_MRI_ was calculated either for both MCAs, *n =* 20, or a single MCA, *n* = 13, based on data quality. All the indices from right and left MCA were averaged for each subject.

*Cardiorespiratory Fitness (CRF):* CRF was estimated based on a non-exercise method developed by [38], using information such as age, gender, body mass index, resting state heart rate and physical activity score. See the *supplementary materials* for physical activity questionnaire and CRF calculation.

*Proposed Index:* The proposed index relies on the information from both forward and reflected waves, however, unlike AI which is amplitude based, the new index only uses the time information. Here, we define the TCD reflection time, *T*_refl_ as the time difference between the occurrence of peak systole and the reflected shoulder of peak systole, in cases of augmented waveforms, and the time difference between two distinct positive peaks, in cases of separated forward and reflected waves, see Fig. 1. Then, we define TCD reflection index, TI_tcd_, as

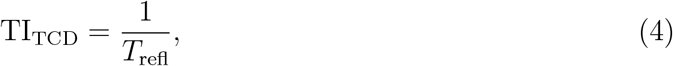

so that TI_TCD_ will be an index of vascular stiffness. TI_TCD_ was calculated for both MCAs using custom Matlab software and then the values were averaged for each subject.

### Statistical Analysis

Statistical analysis was conducted using Matlab^®^. Normality of the data distributions was tested using Lilliefors test with 5% significance level. The correlation coefficients are calculated using Pearson’s correlation for normally distributed data and Spearman’s correlation for non-normal data. Bonferroni corrections with initial α = 0.05 were made for multiple comparisons, resulting in the adjusted level of significance set at 0.0056 for experiment one and 0.0167 for experiment two.

## Results

The correlation matrix for experiment one and experiment two are shown in Table 1. Significant correlations have been marked after adjustment for multiple comparisons. Related scatter plots and least square lines are shown in Fig. 2 and Fig. 3. For D1, two older subjects with high CRF values for their age have been marked by orange arrows (←) in Fig. 2 and Fig. 3. Fig. 2 shows that, unlike AI and PI_TCD_, TI is able to distinguish these subjects from other aged individuals. These cases suggest that TI may better reflect the age-independent impact of CRF on vessel health, which is one of the key motivations for vessel health measures.

**Table 1:** Correlation matrix for experiment one (left) and experiment two (right) reported as *r(p*-value). Correlations found to be significant after Bonferroni correction for multiple comparisons are marked with an asterisk.

|  | Experiment One |  |  |  | Experiment Two |
| --- | --- | --- | --- | --- | --- |
|  | Age | PI<sub>MRI</sub> | CRF |  | Age |
| PI<sub>TCD</sub> | -0.25(0.15) | 0.04(0.87) | 0.10(0.55) | PI<sub>TCD</sub> | 0.44*(0.0009) |
| AI<sub>TCD</sub> | 0.68*(0.44e-5) | 0.38(0.028) | -0.65*(0.15e-4) | AI<sub>TCD</sub> | 0.61*(0.26e-5) |
| TI<sub>TCD</sub> | 0.70*(0.19e-5) | 0.53*(0.002) | -0.79*(0.14e-7) | TI<sub>TCD</sub> | 0.50*(0.0001) |

**Figure 2:**
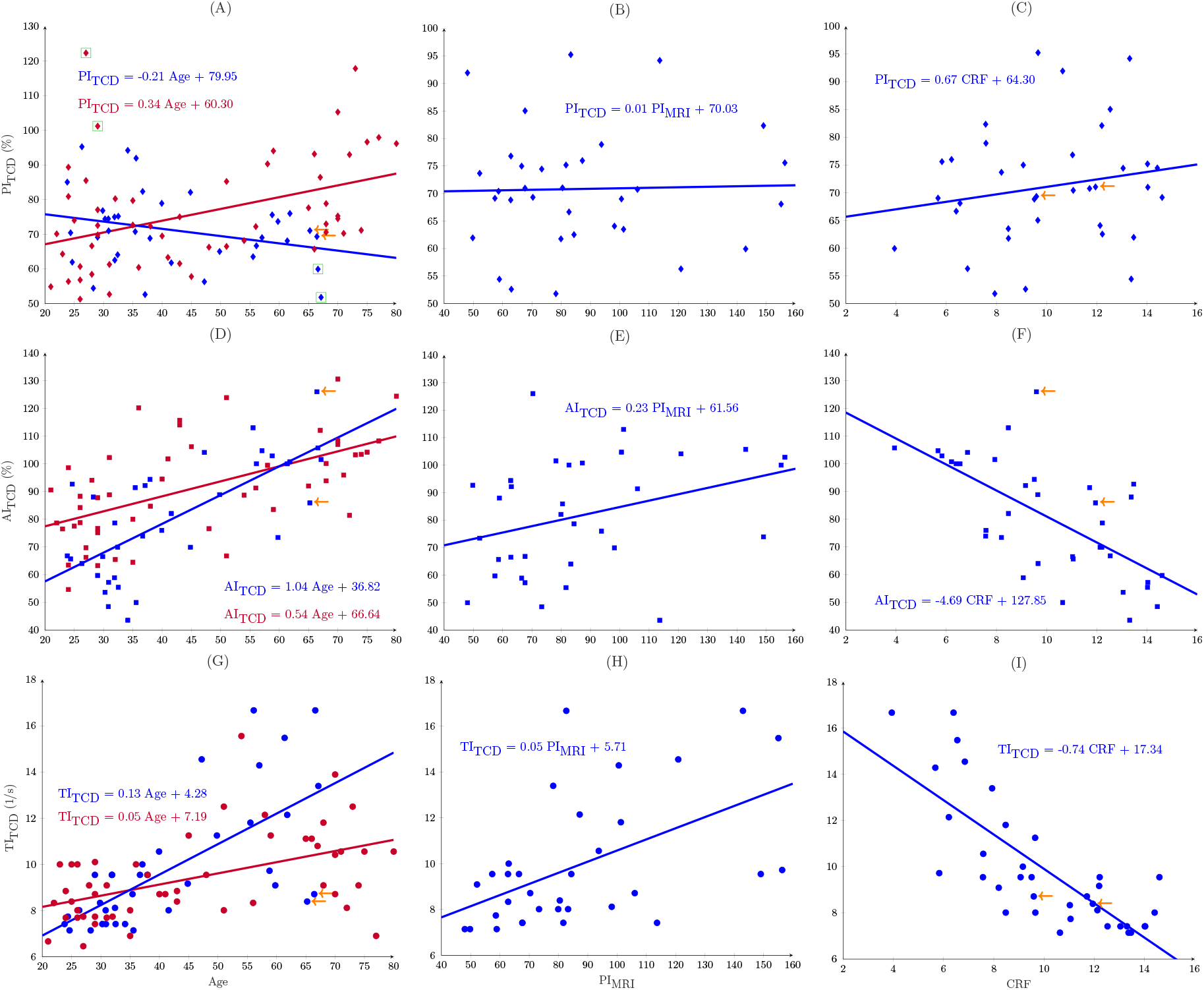
TCD indices plotted against health indices. The lines show least-square fits, D1 is in blue and D2 is in red. Orange arrows track two healthy aged subjects. Green squares are suspected outliers.

**Figure 3:**
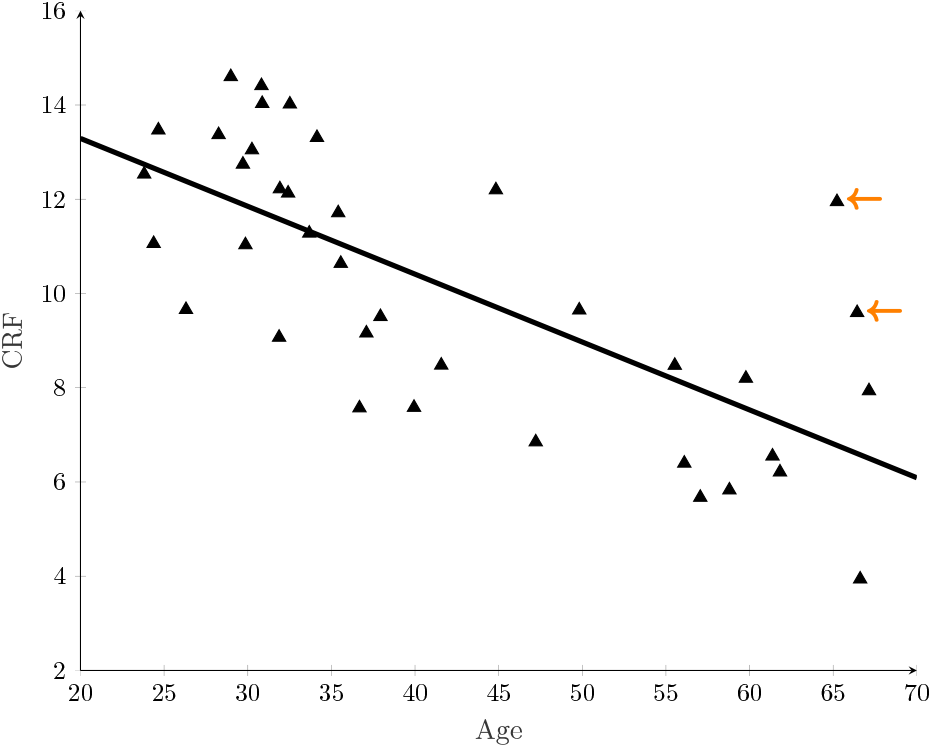
CRF changes with age in D1. Orange arrows show two aged subjects with high CRF values whom are also marked in Fig. 2

In experiment one, PI_TCD_ did not have any significant correlation with age, CRF or PI_MRI_ and the observed weak correlation with age was negative. AI_TCD_ and TI_TCD_ both correlated significantly with age. Although the correlation coefficients of AI_TCD_ and TI_TCD_ are similar with age, TI_TCD_ has higher correlation coefficient with CRF. Among the three Doppler indices only TI_TCD_ has significant correlation with PI_MRI_.

In experiment two, all three Doppler indices, PI_TCD_, AI_TCD_ and TI_TCD_ correlated significantly with age. The correlation coefficient was higher for AI_TCD_ and TI_TCD_ compared to PI_TCD_.

In the scatter plots of Fig. 2.A, there seem to be several data points that can possibly be considered as outliers. These data points have been marked by green squares. The data for these subjects have been carefully re-analyzed, and we have found no reason to remove these subjects. However, if these subjects were removed the correlation coefficients would be *r*_D1_ = −0.12 (not significant) and *r*_D2_ = 0.53 for PI_TCD_ and age.

## Discussion

PI_TCD_ is the most common and easy-to-calculate Doppler index. To understand the controversy in the interpretation of PI_TCD_, it is useful to consider a mathematical model developed in [7] extending the single-element model in [39] to a 3-element Windkessel which describes PI_TCD_ as,

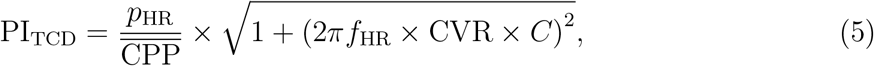

where PI_TCD_ is defined as the fundamental harmonic of the flow velocity divided by the mean flow velocity. *p*_HR_ is the fundamental harmonic of the arterial pulse pressure, *f*_HR_ is the heart rate, *C* is the cerebrovascular compliance and CPP is the cerebral perfusion pressure, the driving source of blood inside the cerebrum [7, 40]. The main point to consider in (5) is that although CVR and *C* affect PI_TCD_ positively, they each change in different directions with age i.e., CVR increases whereas C decreases. For younger adults these effects may cancel out each other hence the lower correlation between PI_TCD_ and both age (*r*_D1_ = −0.25, insignificant) and CRF (*r*_D1_ = 0.10, insignificant). However, for older adults the increased stiffness, or decreased C, may have a dominant effect resulting in a strong decrease in PI_TCD_ even though CVR is increasing. This can explain the low correlations of PI_TCD_ with age for younger adults reported in the literature [34,35]. Our results confirm this for D1, which has a relatively young population, where the correlation between PI_TCD_ and age is negative (r_D1_ = −0.25, insignificant). For D2 (*r*_D2_ = 0.44), PI_TCD_ experiences a bigger increase after the age of 65, as shown in Fig. 2.A.

PI_MRI_ is a different predictor of vascular aging than PI_TCD_ as it considers blood volume changes rather than blood velocity changes. The compliance is the change in blood volume relative to a change in pressure. MRI which measures blood flow, i.e., volume per second, more accurately tracks volume changes than does TCD. If vascular cross-sectional area was known, or if it was constant, it would be possible to derive blood flow from blood velocity. However, changes in vascular cross-sectional area can not be assumed constant as it is actually these changes which derive compliance. In fact, a subtle assumption in deriving (5) is that the vascular cross-sectional area is constant, which is particularly problematic in younger population with compliant arteries. Thus, the uncorrelated PI_TCD_ and PI_MRI_ results (*r*_D1_ = 0.04, insignificant) can be explained by the younger population of D1 and also the lack of cross-sectional area information in PI_TCD_.

AI is an index strongly related to vascular aging factors [41]. Assuming a transmission line with a three-element Windkessel as the load, AI in the pressure waveform can be formulated as [41]

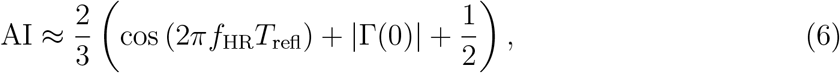

where Γ(0) is called the reflection coefficient and equals the amplitude of the reflected wave divided by the amplitude of the forward wave, measured at the load site [41]. As seen in (6), AI includes T_refl_ and the reflection coefficient, where both are reported to have significant changes with age [29,42]. For AI calculated using the Doppler-based velocity waveform these factors are assumed to contribute in the same way as we have shown a significant correlation of AI_TCD_ with age (*r*_D1_ = 0.68 and *r*_D2_ = 0.61), CRF (*r*_D1_ = −0.65) and PIMRI (*r*_D1_ = 0.38, insignificant). However, heart rate is another contributing factor in AI_TCD_ (see (6)) for which the dependency has also been reported in practice [43]. This is a factor which might change AI independent of vascular compliance or resistance [26].

The timing based index, TI, has a strong relationship with vascular compliance. In the context of the pressure waveform, evidence of change in the time of reflected waves with age have been presented practically [29] and theoretically [41]. As initially suggested in [44] and then mathematically modeled in [41], the time difference between the forward and the backward traveling pressure waves i.e., *T*_refl_, can be given as

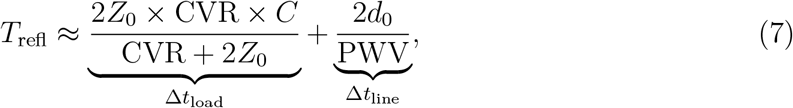

where Δ*t*_line_ is the time delay between the two waves caused by the length of the arteries and Δ*t*_load_ is the delay at the reflection site due to the compliance of the vessels beyond that point and *Z*_0_ is the characteristic impedance. The line delay is inversely proportional to the pulse wave velocity i.e., Δ*t*_line_ ∝ 1/PWV. In addition, the load delay is proportional to the compliance of the vascular bed i.e., Δ*t*_load_ ∝ *C*, and will also decrease as the compliance of the vessels distal to the measurement site reduces with aging. Therefore, similarly we would expect TI_TCD_ to be highly dependent on vascular compliance. The significant correlations of TI_TCD_ with age (*r*_D1_ = 0.70 and *r*_D2_ = 0.50), CRF (*r*_D1_ = −0.79) and with PIMRI (*r*_D1_ = 0.53) support this assumption. We can examine the stronger correlation of TI with CRF than age by considering older subjects with high CRF (see two subjects tracked by arrow signs in Fig. 2, Fig. 3). We hypothesize that the timing index is a predictor of vascular health independent of age, while AI more closely predicts age than CRF. An ideal index of vascular health would not necessarily have a strong correlation with age as lifestyle choices can significantly affect vascular health over time. Specifically, age related progression of aortic and carotid stiffness have been shown to be slower in older participants with a history of high cardiovascular fitness [45].

It should be noted that indices which are influenced by the reflected wave (TI and AI explicitly and PI in some cases) implicitly assume that the distance of travel of the reflected wave is the same across subjects and so that differences in the timing and amplitude of the reflected wave reflect solely the differences in compliance. In the aorta, the gold standard measure of aortic stiffness explicitly estimates the travel distance along the aorta, through external measurement and modeling, and calculates the PWV as the ratio of distance to time difference. This is more difficult to do in the brain.

In general, the application of pressure waveform indices to the Doppler waveform must be done with care due to the differences between the two waveforms. While we often think of a single reflected wave, in reality there are many reflected and re-reflected waveforms adding to form an observed signal. In order to investigate the influence of these factors a simulated flow waveform is shown in Fig. 4 (a simpler form of the ones reported in the literature [46]) which illustrates the underlying reflected and re-reflected waveforms. Reflected flow velocity, i.e. blood which travels toward the heart, is recorded with a negative amplitude by the Doppler device and thus is subtracted from the forward wave velocity. The re-reflected flow, however, is reflected twice during the cardiac cycle and travels in the same direction as the original forward wave. This waveform has a positive amplitude when its velocity is read by the Doppler device. Note that both reflected and re-reflected waveforms have positive amplitudes in pressure readings. Therefore, Doppler-measured velocity waves will have a different shape to pressure waves and the use of the Doppler indices imported from pressure studies must be done with care. However, based on Fig. 4, we can see that an observed reflected peak is more influenced by the re-reflected wave. In other words, the reflected wave plays part in forming a negative dip between the observed forward and reflected peaks and *V*_refl_ is mainly determined by the re-reflected wave. Further, in practice, comparison of simultaneously recorded pressure and flow velocity waveforms has shown that the characteristic time points used in the definitions of AI and TI are similar in both waveforms [47]. This suggests that in both pressure and flow velocity waveforms the reflection time, the pulsatility index and augmentation index account for not just the reflected wave but a combination of reflected and re-reflected waves similarly in both technologies. It also suggests that the stiffness / compliance measured with PI, AI or TI may be equally affected by upstream arteries rather than just the distal cerebral arteries. Given the similarity of timing of the second peak for pressure and flow velocity waveforms we have continued to use this second peak for our derivation of both the AI and TI indices in our calculations. Nonetheless, to our knowledge, there has been no TCD timing index to investigate the effects of the negative reflected wave in the flow velocity waveform and this could be considered in future work.

**Figure 4:**
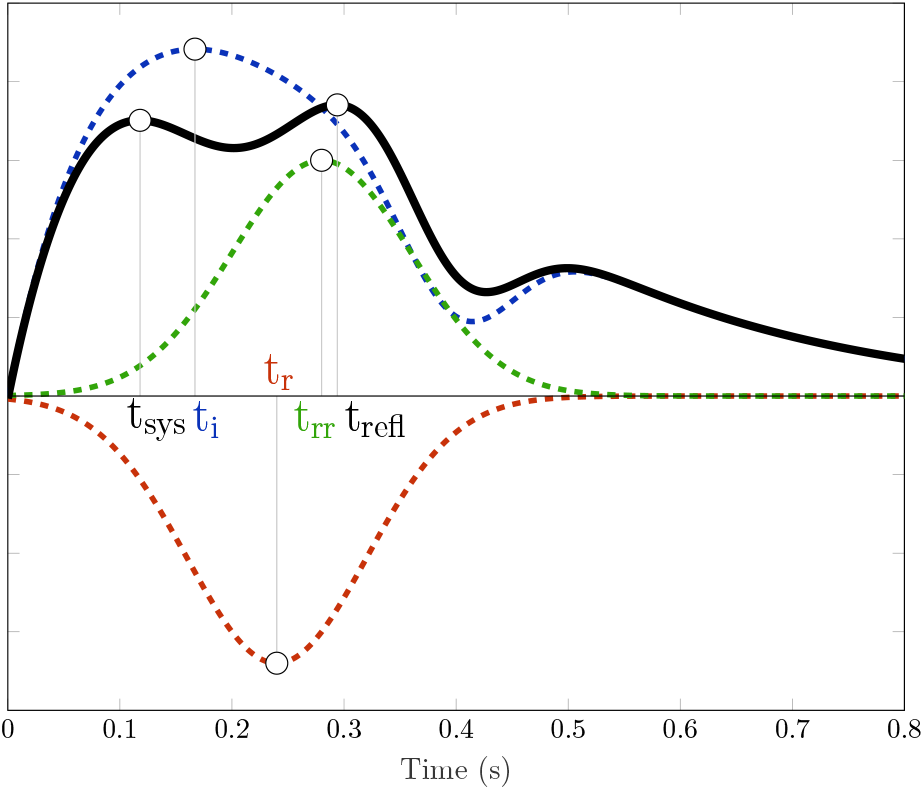
Simulated flow velocity signal to demonstrate summation of incident, reflected and re-reflected waves. Blue, green and red dashed lines represent the incident forward traveling wave, the re-reflected wave and the reflected wave, respectively. Solid line is the sum of the three waveforms. *t*_sys_ and *t*_refl_ are the time of observed systolic and observed reflected peaks. *t_i_*, *t_r_* and *t*_rr_ correspond respectively to time of incident, reflected and re-reflected peaks.

### Limitations

There are several limitations to our study that can potentially change the measured variables due to the study design. The study was conducted on two consecutive days due to the physical location of the MRI and the TCD devices and the two scans were recorded in different positions, i.e. supine and sitting, respectively for MRI and TCD. However, every effort was made to have the scans at the same time of the day and the resting state was established in both sessions by at least 5 minutes sitting during the TCD headpiece set up and at least 5 minutes in supine position in MRI session for acquiring other sequences. In addition, CRF which required heart rate readings was measured on the day of TCD measurement and was not repeated.

## Conclusion

The most commonly used TCD index, PI_TCD_, should be interpreted with caution due to the complex relationships with CVR and vascular stiffness / compliance, each having opposite effects on this index resulting in a weaker correlation between PI_TCD_ and age and no correlation with cardiorespiratory fitness. On the other hand, AI_TCD_ and the new proposed index, TI_TCD_, both showed significant correlations with age linking the two indices to vascular aging. In addition, TI_TCD_ had the strongest correlation with CRF and MRI among the other Doppler indices suggesting that this index may be a suitable replacement for PI_TCD_ and AI_TCD_ when attempting to index cardiovascular health.

## Data Availability

After finalizing the research, de-identified data will be shared with other researchers upon their request.

